# Limited impact of Delta variant’s mutations in the effectiveness of neutralization conferred by natural infection or COVID-19 vaccines in a Latino population

**DOI:** 10.1101/2021.10.25.21265422

**Authors:** Carlos A. Sariol, Crisanta Serrano-Collazo, Edwin J. Ortiz, Petraleigh Pantoja, Lorna Cruz, Teresa Arana, Dianne Atehortua, Christina Pabon-Carrero, Ana M. Espino

**Affiliations:** Department of Microbiology and Medical Zoology, University of Puerto Rico-Medical Sciences Campus, San Juan, PR, USA; Unit of Comparative Medicine, University of Puerto Rico-Medical Sciences Campus, San Juan, PR, USA; Department of Internal medicine, University of Puerto Rico-Medical Sciences Campus, San Juan, PR, USA; Puerto Rico Science, Technology and Research Trust, PR, USA

**Author notes:** Address correspondence to Carlos A. Sariol. These authors have contributed equally to this work and share first authorship (Carlos A. Sariol, Crisanta Serrano-Collazo, Ana M. Espino).

**Keywords:** SARS-CoV-2, COVID-19 Vaccine, Neutralization, Serology, Protection

## Abstract

The SARS-CoV-2 pandemic has impacted public health systems all over the world. The Delta variant seems to possess enhanced transmissibility, but no clear evidence suggests it has increased virulence. Our data shows that pre-exposed individuals had similar neutralizing activity against the authentic COVID-19 strain and the Delta and Epsilon variants. After one vaccine dose, the neutralization capacity expands to all tested variants. Healthy vaccinated individuals showed a limited breadth of neutralization. One vaccine dose induced similar neutralizing antibodies against the Delta compared to the authentic strain. However, even after two doses, this capacity only expanded to the Epsilon variant.

## Background

Coronavirus disease 2019 (COVID-19), caused by the novel severe acute respiratory syndrome coronavirus 2 (SARS-CoV-2), is responsible for the most recent global pandemic declared by the World Health Organization (WHO) on March 11, 2020 [1]. As of October 10, 2021, a total of 6,364,021,792 vaccine doses have been administered worldwide [2]. Despite the tremendous milestone achieved by vaccine approval and administration, SARS-CoV-2, being an RNA virus, has genetically evolved over time leading to the emergence of several variants from different geographic regions [3, 4]. The variant strains have developed characteristics that grant them advantages to maintain viral circulation, such as higher transmissibility and infectivity [5]. Most of these genetic differences are observed in the spike protein (S) region, specifically in the receptor-binding domain (RBD) and the N-terminal domain (NTD). The RBD, and to some extent the NTD as suggested by some evidence, is immunodominant, serving as the main neutralization target by natural and vaccine-elicited antibodies [3, 6]. The Delta variant was first reported in the Indian state of Maharashtra in December 2020 and harbors ten mutations (T19R, G142D, 156del, 157del, R158G, L452R, T478K, D614G, P681R, D950N) in the S protein [3]. Of notice, the Delta variant lacks E484Q, a significant mutation associated with antibody neutralization resistance [7]. After successfully spreading globally, the prevalence of the Delta variant in the USA increased from 1.3% to 94.4% by July 31, 2021 while the Alpha variant decreased from 70% to 2.4% [4]. But perhaps of most serious concern, the Delta variant has been associated with breakthrough infections in vaccinated individuals [4]. The recent surge of cases despite extensive vaccination campaigns supports the concern about low vaccine effectiveness against variants. Studies are at odds regarding this topic, with some claiming that breakthrough infections are more likely to occur due to viral escape from antibodies [8], while others have demonstrated mRNA vaccines remain effective [9]. But still, limited studies are discerning the efficacy of the natural immune response to SARS-CoV-2 vs. the mRNA vaccine-elicited response. Our most recent work confirms that following a natural infection, neutralizing antibody (nAbs) titers generated during infection accompanied by vaccination are significantly better in function than those generated by vaccination alone [10]. To this end, in this study we compared the neutralization capacity of infected vaccinated individuals and healthy vaccinated ones before and after vaccination against several Variants of Concern (VoC) using a surrogate viral neutralization assay [11]. Our results from a Latino population indicate that, compared with vaccination, natural infection induces a broader humoral response offering a wider range of protection against a rapidly evolving virus. These findings have pivotal implications in the understanding of the immune response to COVID-19 induced by vaccination amid emerging variants in the setting of a vaccinated population, and contribute to future vaccine designs and booster schedules.

## Methods

### Study Samples

We selected individuals infected with SARS-CoV-2 any time between March 2020 and February 2021. From 59 subjects followed for months, a subgroup of 10 vaccinated subjects previously exposed to SARS-CoV-2 and a subgroup of 21 healthy-vaccinated volunteers, that were never exposed to SARS-CoV-2, were followed for six to eight months. Vaccinated subjects received either the Pfizer-BioNTech or Moderna vaccine formulations. In the exposed group, all individuals tested positive for SARS-CoV-2 infection by quantitative PCR with reverse transcription (qRT–PCR) or serology tests (IgM and/or IgG). Serum samples from both groups were collected before vaccination (baseline), and after the first and second vaccine doses (Supplementary Tables 1 and 2). Samples used in this study were obtained from adult volunteers (>21 years old) participating in the IRB approved clinical protocol “Molecular Basis and Epidemiology of Viral infections circulating in Puerto Rico”, Pro0004333. Protocol was submitted to, and ethical approval was given by, Advarra IRB on April 21, 2020. Participating volunteers were recruited before the introduction of most of the SARS-CoV-2 variants were reported as circulating in Puerto Rico. More specifically, the Delta variant was first detected on June 15, 2021 [12].

### cPass SARS-CoV-2 Neutralization Antibody Detection Assay

To determine the neutralizing activity of antibodies against SARS-CoV-2, we used a surrogate viral neutralization test (C-Pass GenScript sVNT, Piscataway NJ, USA) according to the manufacturer’s instructions [9-11]. The cutoff for this assay is set to 30% of neutralization. This assay measures the antibodies blocking the RBD-ACE2 interaction and from here, inhibiting viral entry into host cells. For consistency and clarity, the blocking activity is referred to throughout the text as percentage of neutralization.

### Statistical Methods

Statistical analyses were performed using GraphPad Prism 7.0 software (GraphPad Software, San Diego, CA, USA). The statistical significance between or within groups was determined using two-way analysis of variance (ANOVA), one-way ANOVA (Tukey’s, Sidak’s, or Dunnett’s multiple comparisons test as post-hoc test), unpaired t-test, or Wilcoxon–Mann–Whitney, to compare the means. The *p* values are expressed in relational terms with the alpha values. The significance threshold for all analyses was set at 0.05.

## Results

### Natural infection induces an effective neutralization against the Delta variant

To examine the neutralization ability of sera from naturally infected individuals against the Wild Type (WT) SARS-CoV-2, we evaluated baseline samples from 10 volunteers. Out of the 10 subjects, eight had neutralizing activity greater than 70%, indicating the presence of antibodies capable of blocking the RBD-ACE2 binding (Figure 1A and Supplementary Table 3). The other two had neutralization degrees less than 70% but greater than 30%. To compare the neutralizing response elicited by WT SARS-CoV-2 to other virus strains, we exposed sera from those 10 individuals to six variants (Alpha, Beta, Gamma, Epsilon, Kappa and Delta). As expected, the highest neutralizing capacity observed was against the WT strain (Figure 1A). In comparison to the WT strain, there was a significantly decreased neutralizing activity against the Beta, Gamma and Kappa variants (p = 0.0041, p = 0.0003 and p = 0.0294, respectively). Surprisingly, no statistical differences were observed between the WT strain and the Alpha, Epsilon and Delta variants (Figure 1A). These results suggest that natural infection alone is capable of inducing a broad humoral response to various SARS-CoV-2 strains, including the Delta variant.

**Figure 1.**
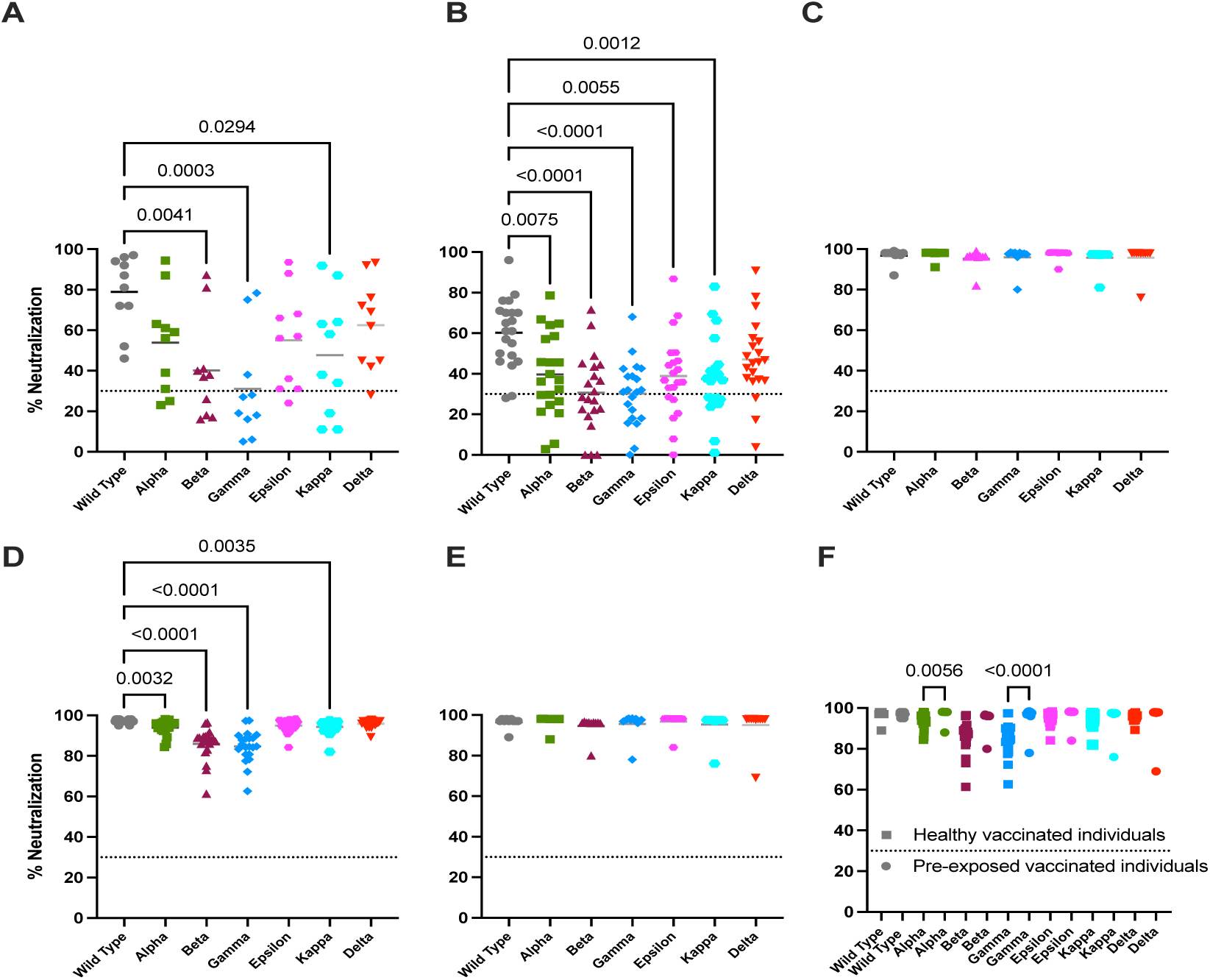
Neutralization capacity of sera from infected and non-infected individuals against SARS-CoV-2 Variants before and after vaccination. The neutralization activity of sera from infected individuals (n=10) and non-infected ones (n=21) before and after vaccination was evaluated against the six variants of concern. Dotted line indicates the limit of detection of the sVNT assay, where the percentage of signal inhibition is determined (≥ 30% indicates a positive result). A Normality test (Shapiro Wilk) was performed for all data sets in order to assess the distribution of the data. The significance threshold for all analyses was set at p<0.05. **A**. Neutralization activity of sera from infected individuals (n=10) before vaccination. A One-Way ANOVA test with Dunnetts’s multiple comparisons test was performed between each of the variants. **B**. Neutralization activity of sera from healthy individuals (n=21) after receiving the 1st vaccine dose. A One-Way ANOVA test with Dunn’s Kruskal-Wallis multiple comparisons test was performed between each of the variants. **C**. Neutralization activity of sera from infected individuals (n=10) after receiving the first vaccine dose. A One-Way ANOVA test with Dunnett’s multiple comparisons test was performed between each of the variants. **D**. Neutralization activity of sera from healthy individuals (n=21) after receiving the 2nd vaccine dose. A One-Way ANOVA test with Dunn’s Kruskal-Wallis multiple comparisons test was performed between each of the variants. **E**. Neutralization activity of sera from infected individuals (n=10) after receiving the 2nd vaccine dose. A One-Way ANOVA test with Dunnett’s multiple comparisons test was performed between each of the variants. **F**. Neutralization activity of sera from vaccinated individuals, pre-exposed (n=10) and healthy (n=21), after receiving the 2nd dose was evaluated. A One-Way ANOVA test with Dunn’s Kruskal-Wallis multiple comparisons test was performed between each of the variants.

### Vaccination boosts neutralizing capacity against variants in previously infected individuals

To assess the humoral immune response to naturally acquired SARS-CoV-2 vs. the mRNA-based COVID-19 vaccine elicited response, we compared the neutralizing capacity of exposed and unexposed subjects after one vaccine dose. Nineteen (19) out of the 21 unexposed individuals (90.5%) produced nAbs (neutralization % >30) (Figure 1B and Supplementary Table 4). Similarly, all previously infected individuals reached neutralizing activity greater than 85% after just one vaccine dose (Figure 1C). This suggests that, in pre-exposed individuals, a single vaccine dose may be sufficient to grant protective immune status against WT SARS-CoV-2. When evaluating the neutralization from unexposed vaccinated individuals against the six VOC, we found significant differences against all except the Delta variant, in comparison with the WT SARS-CoV-2 (p = 0.0075 for Alpha, p < 0.001 for Beta and Gamma, p = 0.0055 for Epsilon and p = 0.0012 for Kappa) (Figure 1B). This suggests that the Delta variant, in our population, does not escape neutralization by antibodies induced by mRNA vaccination. Contrastingly, the neutralization activity in all previously exposed vaccinated individuals increased against all variants with no statistical significant differences (Figure 1C).

### Full vaccination induces limited neutralizing activity against all tested variants in unexposed individuals

Next, we evaluated the neutralizing capacity of antibodies after two vaccine doses in both previously exposed and unexposed individuals. All subjects (n = 31), regardless of immune status before vaccination, reached neutralization levels greater that 95% against WT SARS-CoV-2 after receiving a second vaccine dose (Figures 1D and E). This confirms that, in most COVID-19 naïve individuals, two vaccine doses are required to attain full protection.However, when exploring the neutralization against the variants, unexposed individuals gained similar neutralizing activity to the WT SARS-CoV-2 only against the Epsilon and Delta variants (p = 0.0032 for Alpha, p < 0.001 for Beta and Gamma, and p = 0.0035 for Kappa) (Figure 1D). Therefore, vaccination in unexposed individuals generates a neutralizing response against the Epsilon and Delta variants that is similar to the response against WT SARS-CoV-2 but only after the second dose. Highly relevant, even after the second dose, the neutralization against the other four variants was significantly of lower magnitude compared to the WT.

On the other hand, we observed that the previously infected individuals maintained neutralizing capacity against all variants similar to the response against WT SARS-CoV-2 strain, denoting a key difference in the dynamics of vaccine-elicited antibodies between exposed vs. unexposed individuals (Figure 1E). This difference can be better appreciated in Figure 1F, where both vaccinated groups are compared after receiving the second dose. Of note, neutralization against the Alpha and Gamma variants did not behave similarly between groups, being of higher magnitude in pre-exposed individuals (p = 0.0056 for Alpha and p < 0.0001 for Gamma) (Figure 1F).

## Discussion

There is still very limited information available on the immunity conferred by the natural infection with the authentic SARS-CoV-2 strain or the mRNA COVID-19 vaccines against the viral variants. Using samples collected during the COVID-19 pandemic, most of them before the documented introduction of the variants in the jurisdiction of Puerto Rico [10, 12] we wished to compare the kinetics of the nAbs response in the context of individuals with naturally acquired infection (pre-exposed) and unexposed ones following vaccination via a widely used sVNT [10, 13-15]. Strikingly, we found that natural infection before vaccination confers a broader neutralizing response against different SARS-CoV-2 strains, including the Delta variant, compared to the first dose of the COVID-19 mRNA vaccines. These results are consistent with other reports [16-18] and highlight the need for more epidemiological data about the contribution of previously exposed individuals with natural-acquired immunity to herd immunity. Overall, those subjects are scarcely counted in any statistical model. Highly relevant, our results also suggest that two vaccine doses may induce limited protection against some of the circulating variants in naïve individuals.

Consistent with other works [17, 19, 20] our data confirm that subjects previously exposed to SARS-CoV-2 reach levels of protection just after one vaccine dose against all tested variants. Furthermore, we found a limited contribution, if any, of a second vaccine dose in pre-exposed individuals. Those findings strongly suggest that humoral immunity induced by natural infection results in higher quality antibodies [17, 18, 20] and contributes to the expansion of memory B cells producing more cross-reactive antibodies following vaccination [18]. On the other hand, we found that in naïve subjects, a single dose of the COVID-19 mRNA vaccines induces the same magnitude of nAbs against the Delta variant as to the WT strain. That response is improved after the second dose. However, even after a second dose, the magnitude of neutralization against other variants was significantly lower than that of the WT strain.

A recent remarkable observational study in Puerto Rico collected hospitalization, death, and vaccination rates data for more than 100,000 laboratory-confirmed SARS-CoV-2 infections in a period of 10 months. The study found that the effectiveness of the COVID-19 vaccines preventing hospitalizations or death did not change after the Delta variant became dominant [12]. While that study did not segregate, at an individual level, by the vaccination status of the SARS-CoV-2-positive at the time of hospitalization or death, our results are perfectly aligned and provide the immunological rationale for the findings of that study.

Recent works suggest that the Delta variant may infect vaccinated individuals, defined as breakthrough infections [21]. In vitro neutralization results using monoclonal antibodies argue that vaccination induces a low level of nAbs against the Delta variant [8, 18, 22]. However, as demonstrated by Liu and colleagues, breakthrough infections by the Delta variant may be due to enhanced viral replication and infectivity, and not to antibody evasion or viral immune escape [4]. This statement is reinforced by the fact that the Delta variant lacks the E484Q mutation that seems to grant antibody resistance to other variants [6]. Thus, it looks like that the Delta variant has developed the perfect evolution balance between transmissibility and virulence to become the dominant strain in circulation. However, there is limited or no data from breakthrough infections by the Delta variant in vaccinated people comparing their prior immune status to SARS-CoV-2. Our findings, together with prior reports on the effectiveness of the cellular immune response against the variants [18, 23-25], warrant a revision of COVID-19 vaccine policies implementation in subjects with prior natural immunity to SARS-CoV-2.

We are aware of the limitations of our study, including the small sample size and lack of cellular immunity characterization. The waning of natural or vaccine-elicited immunity remains a possibility outside the follow-up period carried out in this work. However, our results, despite being obtained from a population of different genetic backgrounds, agree with the current ongoing scenario (October 2021) in the United Kingdom (UK). A rampant increase in Delta variant circulation, up 35% over the two previous weeks, has been observed after all restrictions were lifted in summer 2021 [26]. However, taking into account the high number of cases naturally exposed to the virus and a high vaccination rate in the UK [27], as it would be anticipated by our results, the daily deaths are a tenth of what they were in the prior wave [26, 28]. Considering our findings, a more challenging scenario would be a predominance of other variants like Alpha, Beta, Gamma or Kappa, showing limited neutralization after full vaccination with the mRNA COVID-19 vaccines. To our knowledge, this is the first study conducted in a Hispanic/Latino population impacted by COVID-19, and our findings are a significant contribution to the still lacking population-based studies concerning virus-population dynamics in the setting of vaccination and shed light on the design of the second generation COVID-19 vaccines.

## Data Availability

All data produced in the present study are available upon reasonable request to the authors

## Authors Contribution

CAS and AME conceptualized the work and supervised the studies and secured the funds. CSC and PP supervised the work and and supported the figures design. EJO and LC execute the experiments. EJO, CSC, DA and CPC coordinate and supervise the cohort’s management and follow up. EJO, CSC and PP organized the data for future analysis. TA provided administrative and regulatory support. All authors contribute to the results discussion and analysis. CAS and CSC wrote the initial draft, with the other authors providing insights and concepts.

## Acknowledgements

Authors want to thank the volunteers that were willing to participate and contribute to science.

The Puerto Rico Science, Technology and Research Trust supported research reported in this work under agreement number 2020-00272 to AME and CAS. Also, the University of Puerto Rico contributed with the UPR-COVID-19 Grant to CAS and AME. This work was also supported by 1U01CA260541-01 to CAS (NCI/NIAID).

## Corresponding Author contact information

(CAS)

