## Supplementary Table 1 for "Limited impact of Delta variant’s mutations in the effectiveness of neutralization conferred by natural infection or COVID-19 vaccines in a Latino population"

### Supplementary Table 1. Time Between Sample and Vaccination of Healthy and Naturally-Infected Volunteers

| HEALTHY VACCINATED VOLUNTEERS | | | | |
|---|---|---|---|---|
| **Numeric ID** | **Timepoint** | **Time Between First Dose and 1st Sample Post Vac** | **Time Between Second Dose and Sample Post 2nd Vac** | **Time Between First Dose and Last Sample** |
| 479 | Baseline | | | |
| | First Dose | | | |
| 479.2 | Sample P1V | 26 | 16 | 43 |
| | Second Dose | | | |
| 479.3 | Sample P2V | | | |
| 112 | Baseline | | | |
| | First Dose | | | |
| 112.2 | Sample P1V | 21 | 14 | 42 |
| | Second Dose | | | |
| 112.3 | Sample P2V | | | |
| 2 | Baseline | | | |
| | First Dose | | | |
| 2.2 | Sample P1V | 13 | 15 | 36 |
| | Second Dose | | | |
| 2.3 | Sample P2V | | | |
| 3 | Baseline | | | |
| | First Dose | | | |
| 3.2 | Sample P1V | 13 | 15 | 36 |
| | Second Dose | | | |
| 3.3 | Sample P2V | | | |
| 243 | Baseline | | | |
| | First Dose | | | |
| 243.2 | Sample P1V | 15 | 15 | 36 |
| | Second Dose | | | |
| 243.3 | Sample P2V | | | |
| 258 | Baseline | | | |
| | First Dose | | | |
| 258.2 | Sample P1V | 18 | 22 | 43 |
| | Second Dose | | | |
| 258.3 | Sample P2V | | | |
| 119 | Baseline | | | |
| | First Dose | | | |
| 119.2 | Sample P1V | 18 | 18 | 39 |
| | Second Dose | | | |
| 119.3 | Sample P2V | | | |
| 190 | Baseline | | | |
| | First Dose | | | |
| 190.2 | Sample P1V | 12 | 23 | 44 |
| | Second Dose | | | |
| 190.3 | Sample P2V | | | |
| 453 | Baseline | | | |
| | First Dose | | | |
| 453.2 | Sample P1V | 23 | 19 | 42 |
| | Second Dose | | | |
| 453.3 | Sample P2V | | | |
| 6 | Baseline | | | |
| | First Dose | | | |
| 6.2 | Sample P1V | 16 | 14 | 35 |
| | Second Dose | | | |
| 6.3 | Sample P2V | | | |
| 383 | Baseline | | | |
| | First Dose | | | |
| 383.2 | Sample P1V | 23 | 17 | 40 |
| | Second Dose | | | |
| 383.3 | Sample P2V | | | |
| 450 | Baseline | | | |
| | First Dose | | | |
| 450.2 | Sample P1V | 21 | 19 | 40 |
| | Second Dose | | | |
| 450.3 | Sample P2V | | | |
| 110 | Baseline | | | |
| | First Dose | | | |
| 110.2 | Sample P1V | 21 | 19 | 40 |
| | Second Dose | | | |
| 110.3 | Sample P2V | | | |
| 480 | Baseline | | | |
| | First Dose | | | |
| 480.2 | Sample P1V | 21 | 19 | 40 |
| | Second Dose | | | |
| 480.3 | Sample P2V | | | |
| 10 | Baseline | | | |
| | First Dose | | | |
| 10.2 | Sample P1V | 14 | 7 | 28 |
| | Second Dose | | | |
| 10.3 | Sample P2V | | | |
| 116 | Baseline | | | |
| | First Dose | | | |
| 116.2 | Sample P1V | 14 | 7 | 28 |
| | Second Dose | | | |
| 116.3 | Sample P2V | | | |
| 380 | Baseline | | | |
| | First Dose | | | |
| 380.2 | Sample P1V | 14 | 7 | 28 |
| | Second Dose | | | |
| 380.3 | Sample P2V | | | |
| 8 | Baseline | | | |
| | First Dose | | | |
| 8.2 | Sample P1V | 14 | 7 | 28 |
| | Second Dose | | | |
| 8.3 | Sample P2V | | | |
| 117 | Baseline | | | |
| | First Dose | | | |
| 117.2 | Sample P1V | 12 | 6 | 27 |
| | Second Dose | | | |
| 117.3 | Sample P2V | | | |
| 254 | Baseline | | | |
| | First Dose | | | |
| 254.2 | Sample P1V | 12 | 10 | 31 |
| | Second Dose | | | |
| 254.3 | Sample P2V | | | |
| 513 | Baseline | | | |
| | First Dose | | | |
| 513.2 | Sample P1V | 19 | 7 | 35 |
| | Second Dose | | | |
| 513.3 | Sample P2V | | | |
| **AVERAGE Days** | | **17.1** | **14.1** | **36.2** |

| Naturally-Infected Vaccinated Volunteers | | | | |
|---|---|---|---|---|
| **Numeric ID** | **Timepoint** | **Time Between First Dose and 1st Sample Post Vac** | **Time Between Second Dose and Sample Post 2nd Vac** | **Time Between First Dose and Last Sample** |
| 384.2 | Baseline | | | |
| | First Dose | | | |
| 384.3 | Sample P1V | 16 | 23 | 44 |
| | Second Dose | | | |
| 384.4 | Sample P2V | | | |
| 367.2 | Baseline | | | |
| | First Dose | | | |
| 367.3 | Sample P1V | 14 | 20 | 48 |
| | Second Dose | | | |
| 367.4 | Sample P2V | | | |
| 218 | Baseline | | | |
| | First Dose | | | |
| 218.3 | Sample P1V | 28 | 21 | 49 |
| | Second Dose | | | |
| 218.4 | Sample P2V | | | |
| 376.2 | Baseline | | | |
| | First Dose | | | |
| 376.3 | Sample P1V | 19 | 21 | 49 |
| | Second Dose | | | |
| 376.4 | Sample P2V | | | |
| 313.3 | Baseline | | | |
| | First Dose | | | |
| 313.4 | Sample P1V | 17 | 16 | 37 |
| | Second Dose | | | |
| 313.5 | Sample P2V | | | |
| 382.3 | Baseline | | | |
| | First Dose | | | |
| 382.4 | Sample P1V | 12 | 19 | 40 |
| | Second Dose | | | |
| 382.5 | Sample P2V | | | |
| 511 | Baseline | | | |
| | First Dose | | | |
| 511.2 | Sample P1V | 19 | 26 | 47 |
| | Second Dose | | | |
| 511.3 | Sample P2V | | | |
| 512 | Baseline | | | |
| | First Dose | | | |
| 512.2 | Sample P1V | 19 | 26 | 47 |
| | Second Dose | | | |
| 512.3 | Sample P2V | | | |
| 294.2 | Baseline | | | |
| | First Dose | | | |
| 294.3 | Sample P1V | 26 | 32 | 60 |
| | Second Dose | | | |
| 294.4 | Sample P2V | | | |
| 297 | Baseline | | | |
| | First Dose | | | |
| 297.2 | Sample P1V | 19 | 17 | 45 |
| | Second Dose | | | |
| 297.3 | Sample P2V | | | |
| **AVERAGE Days** | | **18.9** | **22.1** | **46.6** |
