## Supplementary Table 2 for "Limited impact of Delta variant’s mutations in the effectiveness of neutralization conferred by natural infection or COVID-19 vaccines in a Latino population"

Supplementary Table 2. Time Between Diagnosis and Vaccines for Pre-exposed Individuals

| ID | Time (days) between Dx and first vaccine dose | Months |
|---|---|---|
| 384 | 111 | 3.7 |
| 367.7 | 90 | 3.0 |
| 218 | 85 | 2.8 |
| 376 | 176 | 5.9 |
| 313 | 169 | 5.6 |
| 382 | 148 | 4.9 |
| 511 | 67 | 2.2 |
| 512 | 67 | 2.2 |
| 294 | 201 | 6.7 |
| 297 | 310 | 10.3 |
| **Average** | **142.4** | **4.7** |

Dx= Diagnosis
