## Supplementary Table 3 for "Limited impact of Delta variant’s mutations in the effectiveness of neutralization conferred by natural infection or COVID-19 vaccines in a Latino population"

### Supplementary Table 3. Neutralization Data Against SARS-CoV-2 and Variants for Pre-exposed Individuals

| ID | Wild Type SARS-CoV-2 | | | V1= (RBD, N501Y) Alpha | | | V2 = (RBD, E484K, K417N, N501Y) Beta | | | V3 = (RBD, E484K, K417T, N501Y) Gamma | | | V4 = (RBD, L452R) Epsilon | | | V5= (RBD, E484Q, L452R) Kappa | | | V6= (RBD, L452R, T478K) Delta | | |
|---|---|---|---|---|---|---|---|---|---|---|---|---|---|---|---|---|---|---|---|---|---|
| | Result (% signal inhibition) | | | | | | | | | | | | | | | | | | | | |
| | Baseline | First Vaccine Dose | Second Vaccine Dose | Baseline | First Vaccine Dose | Second Vaccine Dose | Baseline | First Vaccine Dose | Second Vaccine Dose | Baseline | First Vaccine Dose | Second Vaccine Dose | Baseline | First Vaccine Dose | Second Vaccine Dose | Baseline | First Vaccine Dose | Second Vaccine Dose | Baseline | First Vaccine Dose | Second Vaccine Dose |
| 511 | 81 | 98 | 97 | 59 | 98 | 98 | 41 | 96 | 96 | 27 | 96 | 96 | 56 | 98 | 98 | 58 | 97 | 97 | 62 | 98 | 98 |
| 512 | 46 | 87 | 89 | 25 | 91 | 88 | 16 | 82 | 80 | 5 | 80 | 78 | 31 | 90 | 84 | 19 | 81 | 76 | 45 | 76 | 69 |
| 218 | 72 | 98 | 97 | 31 | 98 | 98 | 26 | 96 | 96 | 6 | 98 | 98 | 31 | 98 | 98 | 11 | 97 | 98 | 45 | 98 | 98 |
| 376 | 94 | 98 | 97 | 63 | 98 | 98 | 38 | 96 | 96 | 28 | 98 | 98 | 68 | 98 | 98 | 63 | 97 | 97 | 72 | 98 | 98 |
| 367 | 92 | 98 | 97 | 56 | 98 | 98 | 40 | 97 | 96 | 38 | 98 | 98 | 66 | 98 | 98 | 64 | 97 | 98 | 76 | 98 | 98 |
| 294 | 52 | 97 | 98 | 23 | 98 | 98 | 17 | 96 | 96 | 19 | 97 | 97 | 24 | 98 | 98 | 11 | 97 | 97 | 28 | 98 | 98 |
| 384 | 87 | 98 | 98 | 61 | 98 | 98 | 37 | 96 | 96 | 18 | 98 | 98 | 57 | 98 | 98 | 38 | 97 | 98 | 69 | 98 | 98 |
| 313 | 97 | 99 | 98 | 87 | 98 | 98 | 81 | 97 | 96 | 75 | 98 | 98 | 88 | 98 | 98 | 87 | 98 | 98 | 92 | 98 | 98 |
| 382 | 72 | 98 | 97 | 39 | 98 | 98 | 18 | 96 | 96 | 16 | 98 | 98 | 36 | 98 | 98 | 34 | 98 | 98 | 42 | 98 | 98 |
| 297 | 96 | 97 | 97 | 94 | 98 | 98 | 87 | 99 | 97 | 78 | 98 | 98 | 94 | 98 | 98 | 92 | 97 | 98 | 93 | 98 | 98 |

POS ≥ 30% signal inhibition

| | |
|---|---|
| **V1**= (RBD, N501Y, Avi & His tag)-HRP | U1100GG280-1:Catalog No: Z03595-100; Name: SARS-CoV-2 Spike protein (RBD, N501Y, Avi & His tag)-HRP; Qty: 1; Size: 100ul |
| **V2** = (RBD, E484K, K417N, N501Y, Avi & His tag)-HRP | U1100GG280-2:Catalog No: Z03596-100; Name: SARS-CoV-2 Spike protein (RBD, E484K, K417N, N501Y, Avi & His tag)-HRP; Qty: 1; Size: 100ul |
| **V3** = (RBD, E484K, K417T, N501Y, Avi & His Tag)-HRP | U1100GG280-3:Catalog No: Z03601-100; Name: SARS-CoV-2 Spike protein (RBD, E484K, K417T, N501Y, Avi & His Tag)-HRP; Qty: 1; Size: 100ul |
| **V4** = (RBD, L452R, Avi & His Tag)-HRP | U1100GG280-4:Catalog No: Z03605-100; Name: SARS-CoV-2 Spike protein (RBD, L452R, Avi & His Tag)-HRP; Qty: 1; Size: 100ul |
| **V5** = (RBD, E484Q, L452R, Avi & His Tag)-HRP | U1100GG280-5:Catalog No: Z03608-100; Name: SARS-CoV-2 Spike protein (RBD, E484Q, L452R, Avi & His Tag)-HRP; Qty: 1; Size: 100ul |
| **V6** = (RBD, L452R, T478K, Avi & His Tag)-HRP | U1100GG280-6Catalog No: Z03614-100; Name: SARS-CoV-2 Spike protein (RBD, L452R, T478K, Avi & His Tag)-HRP; Qty: 1; Size: 100ul |
