## Supplementary Table 4 for "Limited impact of Delta variant’s mutations in the effectiveness of neutralization conferred by natural infection or COVID-19 vaccines in a Latino population"

### Supplementary Table 4. Neutralization Data Against SARS-CoV-2 and Variants for Healthy Vaccinated Individuals

| ID | WT SARS-CoV-2 | | | V1= (RBD, N501Y) Alpha | | | V2 = (RBD, E484K, K417N, N501Y) Beta | | | V3 = (RBD, E484K, K417T, N501Y) Gamma | | | V4 = (RBD, L452R) Epsilon | | | V5= (RBD, E484Q, L452R) India | | | V6= (RBD, L452R, T478K) Delta | | |
|---|---|---|---|---|---|---|---|---|---|---|---|---|---|---|---|---|---|---|---|---|---|
| | Result (% signal inhibition) | | | | | | | | | | | | | | | | | | | | |
| | Baseline | First Vaccine Dose | Second Vaccine Dose | Baseline | First Vaccine Dose | Second Vaccine Dose | Baseline | First Vaccine Dose | Second Vaccine Dose | Baseline | First Vaccine Dose | Second Vaccine Dose | Baseline | First Vaccine Dose | Second Vaccine Dose | Baseline | First Vaccine Dose | Second Vaccine Dose | Baseline | First Vaccine Dose | Second Vaccine Dose |
| 479 | 10 | 46 | 97 | 0 | 26 | 97 | 0 | 0 | 87 | 7 | 31 | 89 | 0 | 33 | 98 | 0 | 42 | 97 | 0 | 43 | 97 |
| 112 | 19 | 96 | 98 | 0 | 79 | 98 | 0 | 72 | 96 | 0 | 68 | 97 | 0 | 87 | 98 | 0 | 83 | 97 | 0 | 91 | 97 |
| 2 | 10 | 46 | 98 | 3 | 21 | 95 | 0 | 23 | 90 | 5 | 43 | 90 | 0 | 29 | 97 | 0 | 26 | 95 | 0 | 28 | 96 |
| 3 | 19 | 49 | 98 | 2 | 37 | 94 | 0 | 14 | 86 | 0 | 22 | 83 | 0 | 50 | 96 | 0 | 25 | 96 | 0 | 45 | 97 |
| 243 | 22 | 76 | 97 | 0 | 46 | 96 | 0 | 27 | 89 | 6 | 32 | 90 | 2 | 42 | 97 | 0 | 36 | 96 | 0 | 56 | 97 |
| 258 | 20 | 50 | 97 | 0 | 32 | 94 | 0 | 23 | 73 | 0 | 18 | 81 | 0 | 20 | 94 | 0 | 24 | 93 | 0 | 37 | 95 |
| 119 | 21 | 29 | 97 | 0 | 5 | 86 | 0 | 0 | 61 | 0 | 3 | 63 | 0 | 8 | 93 | 0 | 1 | 91 | 0 | 17 | 94 |
| 190 | 23 | 28 | 97 | 0 | 3 | 96 | 0 | 0 | 88 | 4 | 0 | 86 | 0 | 0 | 94 | 0 | 7 | 94 | 0 | 4 | 96 |
| 453 | 12 | 76 | 97 | 1 | 64 | 93 | 0 | 35 | 83 | 0 | 29 | 84 | 0 | 65 | 94 | 0 | 66 | 94 | 0 | 78 | 96 |
| 6 | 15 | 65 | 98 | 5 | 67 | 96 | 1 | 45 | 88 | 1 | 31 | 78 | 0 | 36 | 91 | 0 | 41 | 91 | 0 | 58 | 94 |
| 383 | 4 | 71 | 98 | 2 | 59 | 96 | 0 | 44 | 90 | 4 | 51 | 91 | 0 | 44 | 97 | 0 | 58 | 96 | 0 | 63 | 97 |
| 450 | 4 | 70 | 95 | 0 | 57 | 84 | 1 | 37 | 75 | 0 | 39 | 72 | 8 | 40 | 84 | 0 | 38 | 82 | 0 | 53 | 89 |
| 110 | 18 | 44 | 97 | 4 | 36 | 92 | 0 | 29 | 89 | 2 | 15 | 88 | 0 | 27 | 92 | 0 | 28 | 92 | 0 | 38 | 94 |
| 480 | 4 | 70 | 95 | 0 | 25 | 95 | 11 | 22 | 82 | 0 | 25 | 81 | 0 | 18 | 95 | 2 | 27 | 94 | 0 | 36 | 96 |
| 10 | 24 | 61 | 97 | 0 | 40 | 96 | 0 | 19 | 89 | 0 | 16 | 85 | 1 | 33 | 96 | 0 | 27 | 96 | 0 | 37 | 97 |
| 116 | 0 | 66 | 97 | 0 | 46 | 97 | 0 | 39 | 92 | 0 | 37 | 89 | 0 | 46 | 97 | 5 | 40 | 96 | 0 | 50 | 97 |
| 380 | 25 | 61 | 97 | 0 | 30 | 94 | 0 | 49 | 88 | 3 | 43 | 84 | 0 | 50 | 95 | 1 | 39 | 95 | 0 | 49 | 96 |
| 8 | 26 | 70 | 97 | 2 | 46 | 92 | 0 | 44 | 87 | 0 | 39 | 85 | 6 | 37 | 95 | 0 | 28 | 95 | 0 | 46 | 96 |
| 117 | 22 | 55 | 97 | 0 | 21 | 89 | 0 | 31 | 86 | 0 | 18 | 78 | 0 | 36 | 95 | 0 | 45 | 95 | 6 | 41 | 96 |
| 254 | 18 | 57 | 98 | 2 | 30 | 96 | 0 | 28 | 90 | 0 | 32 | 89 | 13 | 45 | 98 | 0 | 37 | 97 | 0 | 47 | 97 |
| 513 | 17 | 79 | 97 | 5 | 65 | 98 | 0 | 64 | 97 | 2 | 42 | 97 | 17 | 69 | 98 | 1 | 69 | 98 | 0 | 73 | 98 |

**POS ≥ 30% signal inhibition**

| | |
|---|---|
| **V1** = (RBD, N501Y, Avi & His tag)-HRP | U1100GG280-1:Catalog No: Z03595-100; Name: SARS-CoV-2 Spike protein (RBD, N501Y, Avi & His tag)-HRP; Qty: 1; Size: 100ul |
| **V2** = (RBD, E484K, K417N, N501Y, Avi & His tag)-HRP | U1100GG280-2:Catalog No: Z03596-100; Name: SARS-CoV-2 Spike protein (RBD, E484K, K417N, N501Y, Avi & His tag)-HRP; Qty: 1; Size: 100ul |
| **V3** = (RBD, E484K, K417T, N501Y, Avi & His Tag)-HRP | U1100GG280-3:Catalog No: Z03601-100; Name: SARS-CoV-2 Spike protein (RBD, E484K, K417T, N501Y, Avi & His Tag)-HRP; Qty: 1; Size: 100ul |
| **V4** = (RBD, L452R, Avi & His Tag)-HRP | U1100GG280-4:Catalog No: Z03605-100; Name: SARS-CoV-2 Spike protein (RBD, L452R, Avi & His Tag)-HRP; Qty: 1; Size: 100ul |
| **V5** = (RBD, E484Q, L452R, Avi & His Tag)-HRP | U1100GG280-5:Catalog No: Z03608-100; Name: SARS-CoV-2 Spike protein (RBD, E484Q, L452R, Avi & His Tag)-HRP; Qty: 1; Size: 100ul |
| **V6** = (RBD, L452R, T478K, Avi & His Tag)-HRP | U1100GG280-6Catalog No: Z03614-100; Name: SARS-CoV-2 Spike protein (RBD, L452R, T478K, Avi & His Tag)-HRP; Qty: 1; Size: 100ul |
